# A cross sectional study on “Behavioral risk factors of cardiovascular disease among adolescents of secondary level school in a sub-metropolitan city of Nepal”

**DOI:** 10.1101/2024.05.10.24307183

**Authors:** Sita Bista, Bishow Puri, Sanju Maharjan, Buna Bhandari

## Abstract

**Background:** Cardiovascular diseases (CVDs) are the leading cause of death and disability globally, where one-third of adolescents are affected. There is plenty of evidence that behavioral factors like physical inactivity, unhealthy diet, smoking, and alcohol use are major risks associated with CVDs among adolescents/school-going children.

**Objective(s):** The main aim of this study was to assess the prevalence of behavioral risk factors of cardiovascular disease among adolescents of secondary-level schools in Tulsipur Sub Metropolitan City, Nepal.

**Methods:** It was a school-based cross-sectional study among 361 adolescents of grades 11 and 12 between the ages of 16 and 19. The school was selected by using a stratified proportionate sampling method. Data were collected through a self-administered structured and validated questionnaire containing socio-demographic characteristics, behavioral risk factors of CVDs and respective parent’s information. Data were analyzed using both descriptive and analytical statistics.

**Results:** The most prevalent behavioral risk factor was consumption of calorie drinks (99%); followed by sedentary behavior (60%), insufficient fruit and vegetable intake (57%), physical inactivity (35%), intake of processed food high in salt (33%), refined vegetable oil use in meal preparation (19%) and added salt intake (15.5%). Similarly, the prevalence of current smoking tobacco, alcohol use and smokeless tobacco use was 12%, 10% and 9%, respectively. Mother education, ethnicity, education system were associated factors of majority of behavioral risk factors of CVDs among adolescents. Mother education, parent’s smoking habit and parent’s smokeless tobacco use were associated factors of current tobacco smoking. Parent’s smokeless tobacco use and parent’s alcohol use were associated factors of current alcohol use.

**Conclusions:** This study provided evidence of high prevalence of CVD risk factors among adolescents in Nepal. It demands an urgent need to effectively design appropriate interventions in household and school settings to address the risk factors at the municipal level of Nepal.

## INTRODUCTION

Cardiovascular disease (CVD) is the leading cause of mortality with 17.9 million deaths globally which attributed to 32% of all deaths in 2019 (1). The burden is high especially at low and middle income countries (LMICs) that accounts for a three quarters of deaths (1). It is believed that approximately 23 million individuals will die from CVD by 2030 if current trends persist (2). CVD affects one third of young school going children globally, making it the largest epidemic to human population (3). It is one of the most significant public health challenges faced by adolescents, aged 10-24 years that accounts for over 25% of global population. In 2017, CVDs contributed to 26·9% of total deaths and 12·8% of total DALYs in Nepal (4). Surprisingly, deaths from CVD among 15-19 age group was 3.9 deaths per 100,000 that accounts for 4.8% of total CVD deaths in Nepal (4). This has the potential of creating a huge burden on macroeconomic conditions as CVD mainly occurs in productive age (1).

There are plenty of evidence that physical inactivity, unhealthy diet, smoking, and alcohol use are major risks associated with CVDs among adolescents and school going children (3). These risk factors starts in childhood, accelerate during adolescents though early clinical indications of CVD are usually visible in adulthood (2). LMICs including Nepal are going through transition of rapid unplanned urbanization. Children living in urban areas do not have adequate space for physical activity. In addition, they are exposed with access to junks and fast foods, making them vulnerable to negative health impacts including risk of CVDs (5). There is evidence that 12.1 % of adolescents used tobacco, 15.7% of them used alcohol, 74.3% of them had low fruit and vegetable intake and 71.4 % reported low physical activity between 2003 and 2011 in LMICs setting (6). These figures are alarming and need immediate attention especially in this age group to combat CVD in LMICs such as in Nepal. However, there are limited evidence on CVD risk factors in the semi urban area of Nepal. Therefore, this study was conducted to assess the behavioral risk factors of CVDs among adolescents of the semi-urban area of Nepal.

## METHODS

### Study design and setting

A cross sectional study was conducted among secondary level school going adolescents of age between 16-19 years, in Tulsipur Sub-Metropolitan City, Nepal. Tulsipur Sub-Metropolitan City is in the mid-western region of Nepal, where rapid urbanization has occurred as people from neighboring districts descend for better education and opportunities. Provincial capitals being situated in the district, opening of fast-food chains, changing lifestyles, and feeding habits pose a risk of CVDS.

### Study population

Students of grade 11 and 12 studying in both public and private schools between the age group of 16-19 years were included in the study. The students with a history of CVDs and other chronic diseases were excluded.

### Operational Definition of Key Variables Of The Study

#### Current smoker/ smokeless tobacco user/ alcohol user

Those who are using the products in past 30 days preceding the study (7).

#### Inadequate fruits and/ or vegetables intake

Less than five servings of fruits and/or vegetable a day was considered as inadequate (7).

#### Added salt intake

It was defined for study participants who identified to add dietary salt to food always or often during eating (7).

#### Consumption of processed food high in salt

Processed food high in salt means foods that has been altered from their natural state, such as packaged salty locally available snacks canned salty food, salty food prepared at fast food restaurant, cheese, processed meat, etc. Consumption of processed food high in salt was defined for those participants who used to consume processed food high in salt always or often (7).

#### Adequate physical activity

Adequate physical activity was defined as moderate to vigorous activity greater than or equal to 60 min/day for all 7 days of previous week (7).

#### Sedentary behavior

Sedentary behavior included spending time in school, watching television and mobiles and playing video games. Spending time only by sitting or lying down for 9 hours or more, per day as defined as sedentary behavior (8).

### Data Sources

Sampling frame was obtained from the education section of the sub-metropolitan city and from administration section of respective schools.

### Bias

A self-administered questionnaire was used to assess the risk factors that might lead to self-reported and recall bias. Collecting data in a group setting might have been subjected to information bias which was addressed by not allowing more than 15 participants with adequate spacing. However, we have used standardized questions with appropriate probing. In addition, enough clarification of questions was provided before data collection to overcome those biases.

#### Sample size

The total calculated sample was 367 with the assumptions [Number of secondary level school adolescents in Tulsipur Sub-Metropolitan City(N) = 5262 enlisted in sampling frame provided by education section of Tulsipur Sub-Metropolitan City, level of confidence at α= 0.05, Z= 1.96 at 95% of confidence interval, p= estimated proportion in population=0.366 (prevalence of currently using any tobacco products in Lumbini Province according to STEPs survey Nepal 2019, 10% non-response rate]

#### Sampling

Stratified proportionate sampling method was used. First, the two strata were two secondary public and private schools. The acquired sample size was proportionately distributed to each stratum based on population distribution. Two secondary public schools and two secondary private schools were selected at random. Among selected schools, students were enrolled in study through simple random (lottery) method using random number generator. The students were from different streams which were further divided into grades 11 and 12 with a proportional distribution belonging to age group 16-19. Detailed sampling frame is depicted in fig 1

**Figure 1.**
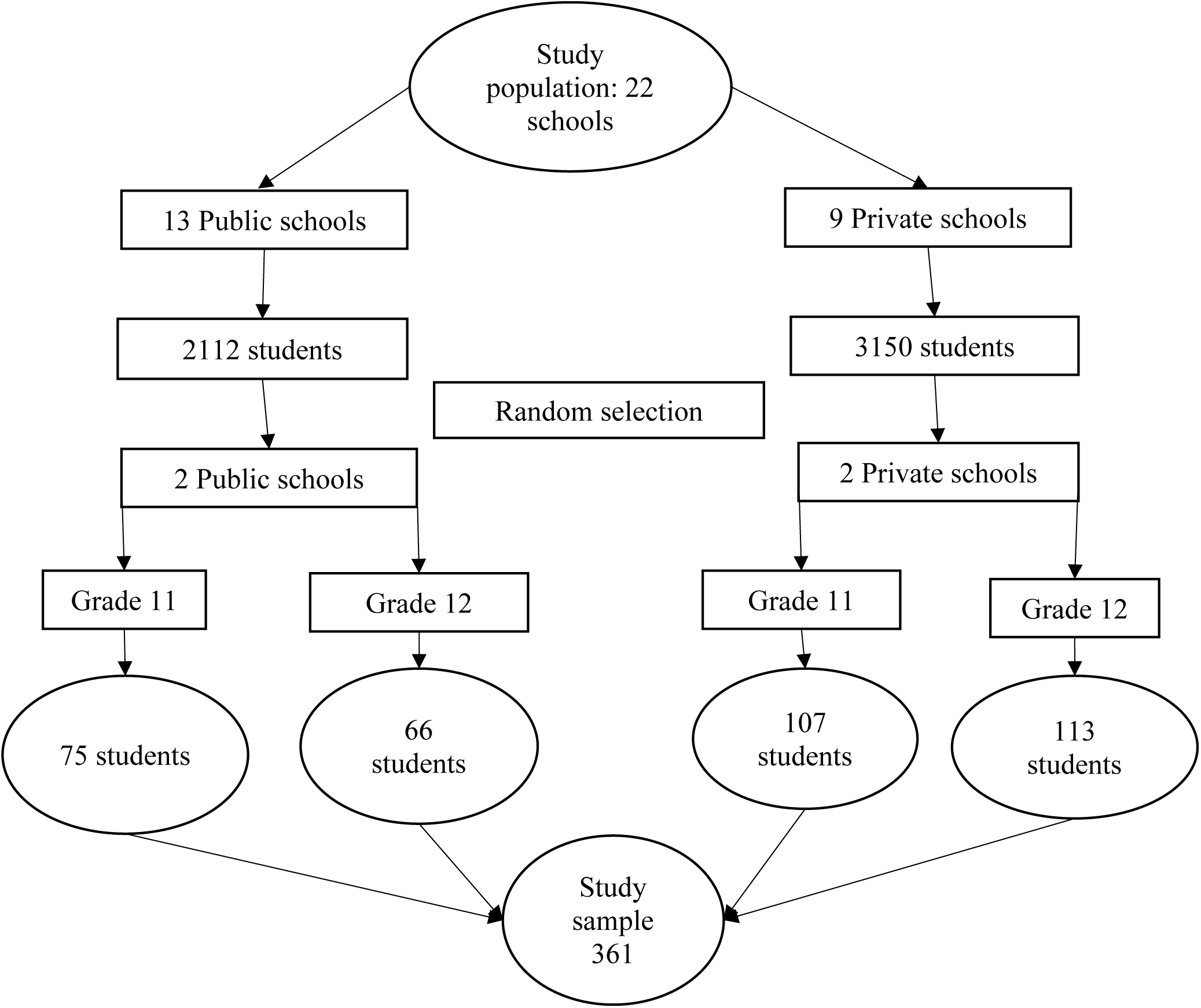
Sampling design.

### Data collection tools and techniques

Data was collected using self-administered Nepali questionnaire after obtaining written consent from parents and verbal consent from students. The structured questionnaire was adopted from a study conducted by Islam.et.al among school going children in Bangladesh after permission was obtained (9). Which contains 18 screening questions, covering all the five CVD risk behaviors. The questionnaire was translated into Nepali and pretested after among 10 % of sample size in similar settings and revised as per needed. During data collection, strict supervision was done to minimize information bias by not keeping more than 15 participants in a room. The questions were clearly addressed and explained before data collection.

### Data processing and analysis

All collected information was systematically compiled, coded, checked, and edited on the same day of data collection. Obtained data was entered in Epi-data version 4.6 and rechecked and cleaned after entry, to ensure the quality of data. Analysis was done using SPSS version 22.0. Univariate analysis was done by calculating frequencies, percentages, mean and median for social and demographic variables of respondents and parent’s information. Bivariate analysis was done using chi-square test for association of socio-demographic characteristics and parents’ information with behavioral risk factors of CVDs at the level of 95% of confidence level.

### Ethical considerations

Ethics approval was taken from the Institutional Review Committee of Tribhuvan University Institute of Medicine, Nepal [IRC no: 355(6–11) E2 078/079]. The consent of respective parents of participants and permission of school authority was taken. Informed written consent was obtained from the respondents and parents, respectively. Anonymity and confidentiality were maintained by not sharing individually identifiable information.

## RESULTS

A total of 367 participants filled in the self-administered questionnaire but six responses needed to be completed. Hence the final analysis comprised 361 responses.

### Sociodemographic characteristics of the school children

The mean ± SD age of the participants was 17.5±0.92. More than half (54 %) of the participants were female and 52.6% belonged to Chhetri ethnicity. Around half of them (50.4%) were studying grade 11 and 61% from management faculty. Around three fourth (66.8 %) of them were from private education system and 76% were living with their family members.

**Table 1.** Social and demographical characteristics of the participants.

| Socio-demographic characteristics | Number | Percentage |
| --- | --- | --- |
| **Age** |  |  |
| 16 | 60 | 16.6% |
| 17 | 113 | 31.3% |
| 18 | 143 | 39.6% |
| 19 | 45 | 12.5% |
| Mean ± SD | | ?? |
| **Gender** |  |  |
| Male | 164 | 45.4 |
| Female | 195 | 54.0 |
| Don't want to disclose | 2 | 0.6 |
| **Ethnicity** |  |  |
| Chhetri | 190 | 52.6 |
| Janajati | 68 | 18.8 |
| Brahmin | 65 | 18.0 |
| Dalit | 32 | 8.9 |
| Muslim | 2 | 0.6 |
| Others | 4 | 1.1 |
| **Religion** |  |  |
| Hindu | 354 | 98.1 |
| Christian | 4 | 1.1 |
| Muslim | 2 | 0.6 |
| Budhism | 1 | 0.3 |
| **Grade** |  |  |
| 11 | 182 | 50.4 |
| 12 | 179 | 49.6 |
| **Education System** |  |  |
| Private | 241 | 66.8 |
| Public | 120 | 33.2 |
| **Current living condition** |  |  |
| Living with family members | 274 | 76 |
| Renting a room with friend | 61 | 16.9 |
| Alone | 21 | 5.8 |
| Others | 5 | 1.4 |

### Socio-demographic, medical and behavioral characteristics of parents

Around one quarter (24 %) of respondent’s mother and 10.5% of respondent’s father had no formal schooling. The majority of respondent’s father (29%) were involved in business whereas 63 % of respondent’s mother were homemakers. Overall, 20 % of them reported their parents have chronic illness, 15% were smoker, 24% consume smokeless tobacco and 27% consume alcohol.

**Table 2:** Socio-demographical and medical characteristics of parents.

| Characteristics | Numbers | Percentage |
| --- | --- | --- |
| **Mother education** | **n= 357\*** |  |
| No formal schooling | 90 | 24.9 |
| Less than primary school | 67 | 18.6 |
| Primary school | 79 | 21.9 |
| Secondary school | 60 | 16.6 |
| High School | 41 | 11.4 |
| College/University | 18 | 5.0 |
| Post graduate degree | 2 | 0.6 |
| **Mother occupation** | **n=357\*** |  |
| Home maker | 229 | 63.4 |
| Agriculture | 47 | 13.0 |
| Business | 39 | 10.8 |
| Government employee | 23 | 6.4 |
| Non-government employee | 8 | 2.2 |
| Foreign employment | 6 | 1.7 |
| Unemployed | 4 | 1.1 |
| Retired | 1 | 0.3 |
| **Father education** | **n=350\*\*** |  |
| No formal education | 38 | 10.5 |
| Less than primary school | 59 | 16.3 |
| Primary School | 47 | 13.0 |
| Secondary school | 101 | 28.0 |
| High School | 67 | 18.6 |
| College/University | 26 | 7.2 |
| Post graduate degree | 13 | 3.6 |
| **Father occupation** | **n=350 \*\*** |  |
| Business | 105 | 29.1 |
| Agriculture | 101 | 28.0 |
| Foreign employment | 41 | 11.4 |
| Non-government employee | 40 | 11.1 |
| Government employee | 32 | 8.9 |
| Home-maker | 15 | 4.2 |
| Unemployed | 12 | 3.3 |
| Retired | 4 | 1.1 |
| **Chronic illness of father or mother or both** | **n= 358\*\*\*** | *\*multiple response* |
| Hypertension | 48 | 13.4 |
| Heart Disease | 10 | 2.8 |
| Diabetes mellitus | 11 | 3 |
| Cancer | 1 | 0.3 |
| Chronic respiratory | 4 | 1.1 |
| **Behavioral risks of father or mother or both** | **n=358\*\*\*** | *\*multiple response* |
| Smoking habit | 53 | 14.8 |
| Smokeless tobacco | 88 | 24.4 |
| Alcohol use | 98 | 27.4 |

\*4 children don't have mother
\*\*11 children don't have father
\*\*\* 3 children neither have father nor mother

### Distribution of CVD risk factors among the participants

The most prevalent behavioral risk factor was the consumption of calorie drinks (99%), followed by sedentary behavior (60%). The third most prominent behavioral risk for CVD was insufficient fruit and vegetable intake (57%), followed by physical inactivity (35%), intake of processed food high in salt (33%), refined vegetable oil use in meal preparation (19%), added salt intake (15.5%). Similarly, the prevalence of current smoking tobacco, alcohol use and smokeless tobacco use was 12%, 10%and 9%, respectively, as presented in table 3.

**Table 3.** : Prevalence of behavioral risk factors among secondary school level adolescents n=361.

| Behavioral risk factor | Number | Percentage (%) |
| --- | --- | --- |
| Calorie drink consumption | 356 | 99 |
| Sedentary behavior (Sitting or lying down for 9 hrs. or more) | 217 | 60 |
| Insufficient fruit and vegetable intake (less than 5 servings per day) | 206 | 57 |
| Physical inactivity | 126 | 34 |
| Processed food high in salt intake (Always or often) | 119 | 33 |
| Refined vegetable oil use (cooking) | 70 | 19 |
| Salt addition in food (Always or often) | 56 | 15.5 |
| Current smoking tobacco use | 44 | 12 |
| Current alcohol user | 37 | 10 |
| Current smokeless tobacco use | 31 | 9 |

**Table 4:**
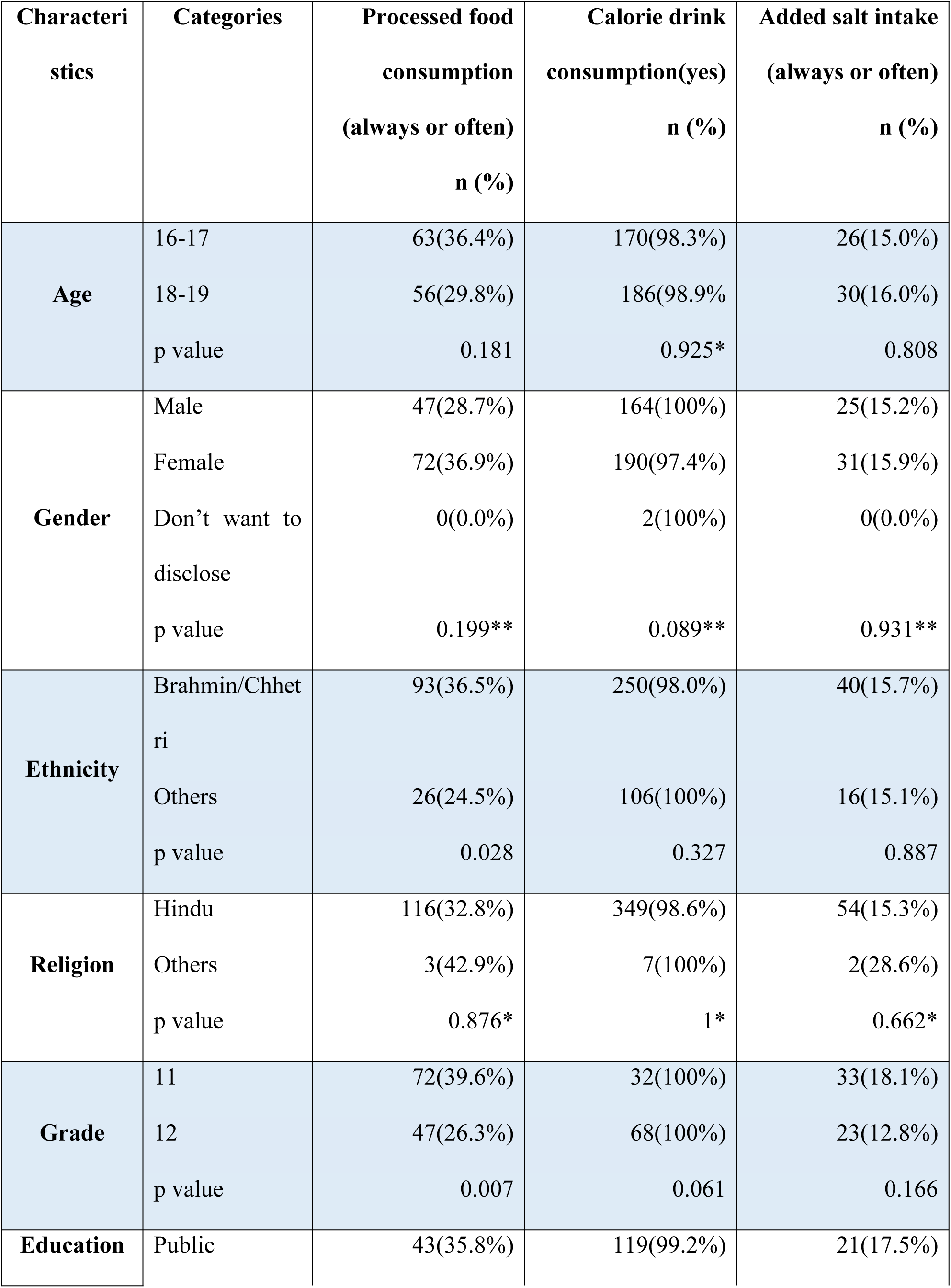
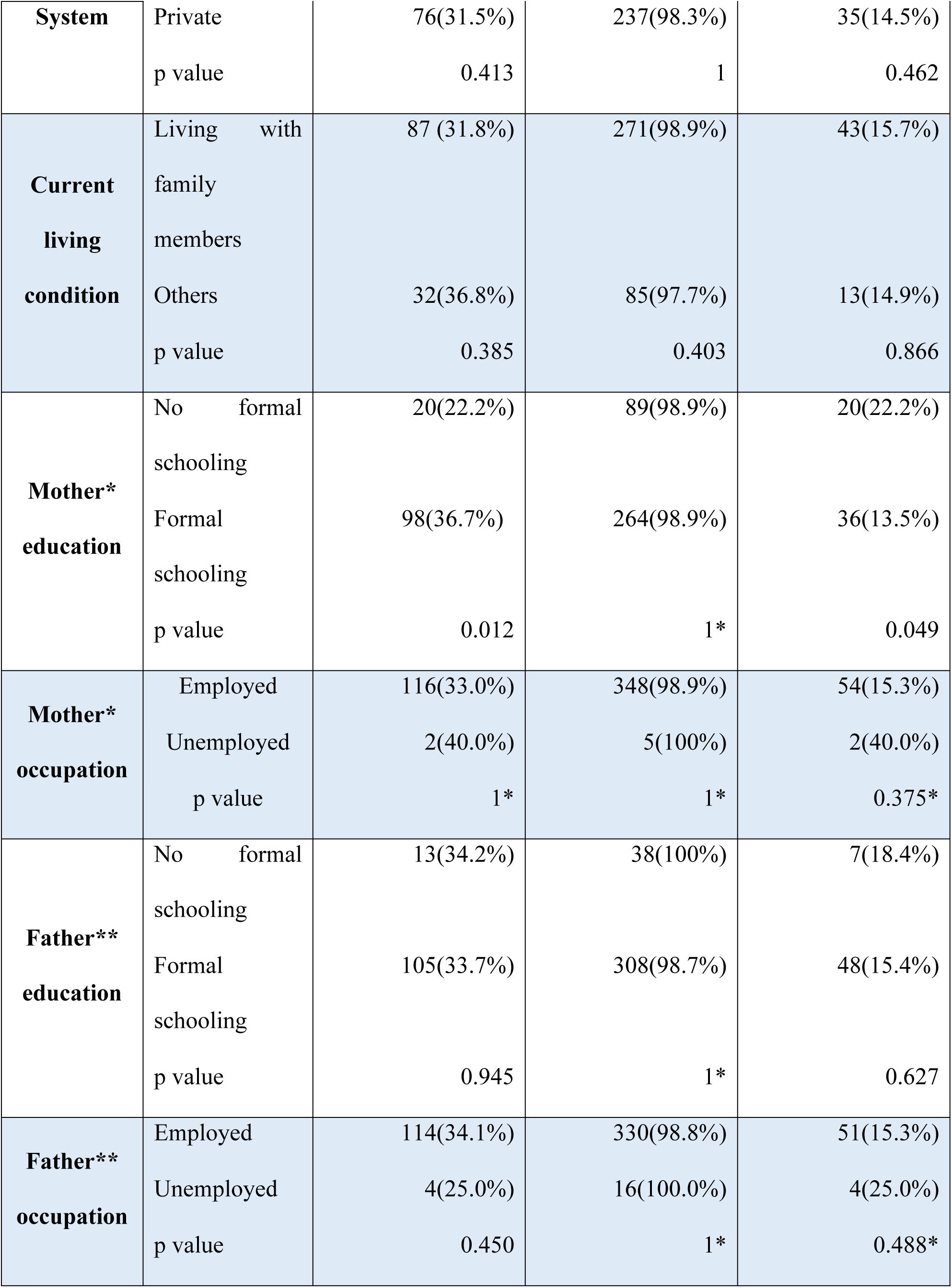
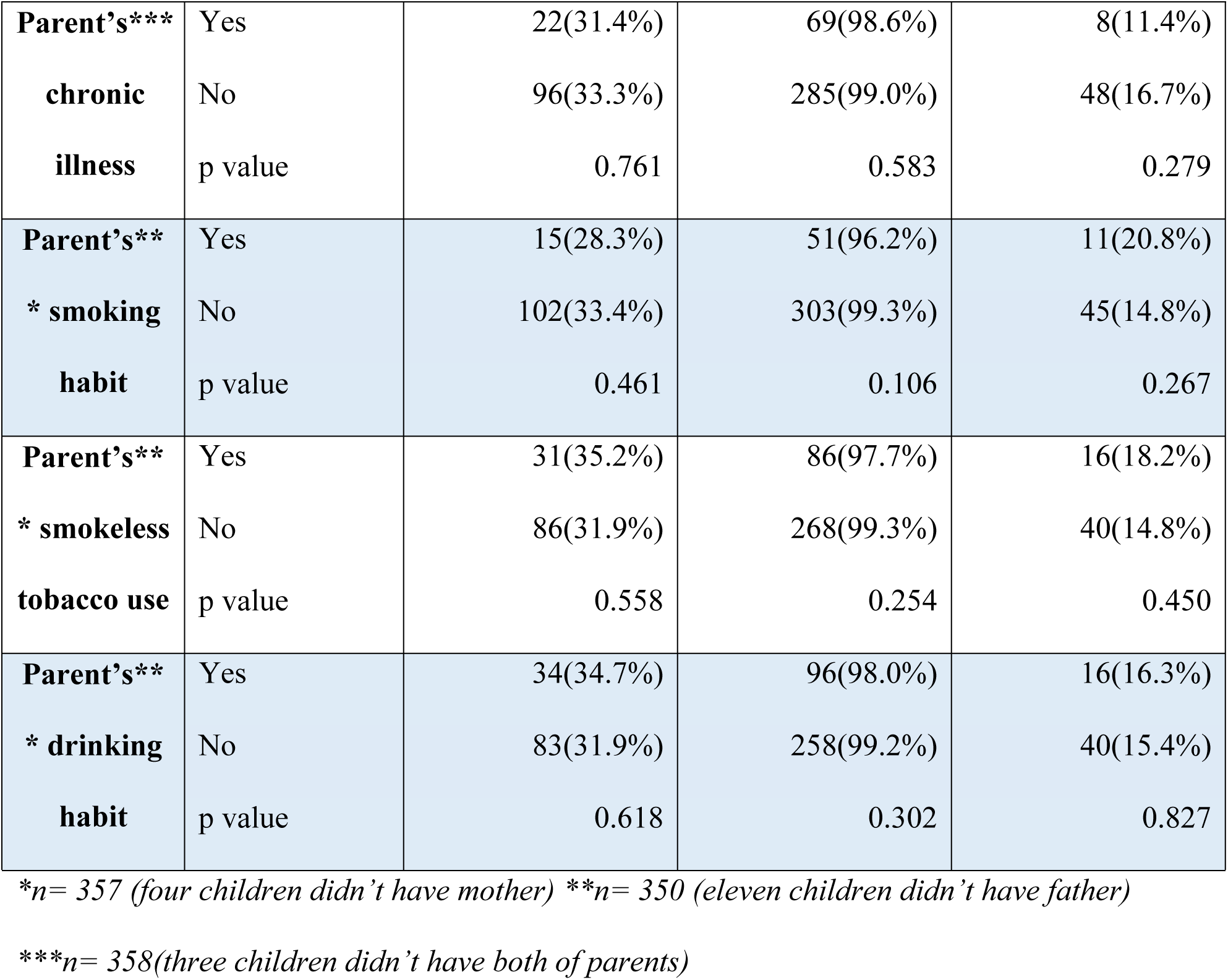
Association of processed food high in salt consumption, calorie drink consumption and added salt intake.

Further, sedentary behavior was associated with education system (p = 0.003) and physical inactivity was associated with education system (p <0.001), mother’s education (0.025), and father education (p = 0.046). Insufficient fruit and vegetable intake was not associated with any of the variables as presented in table 5.

**Table 5:**
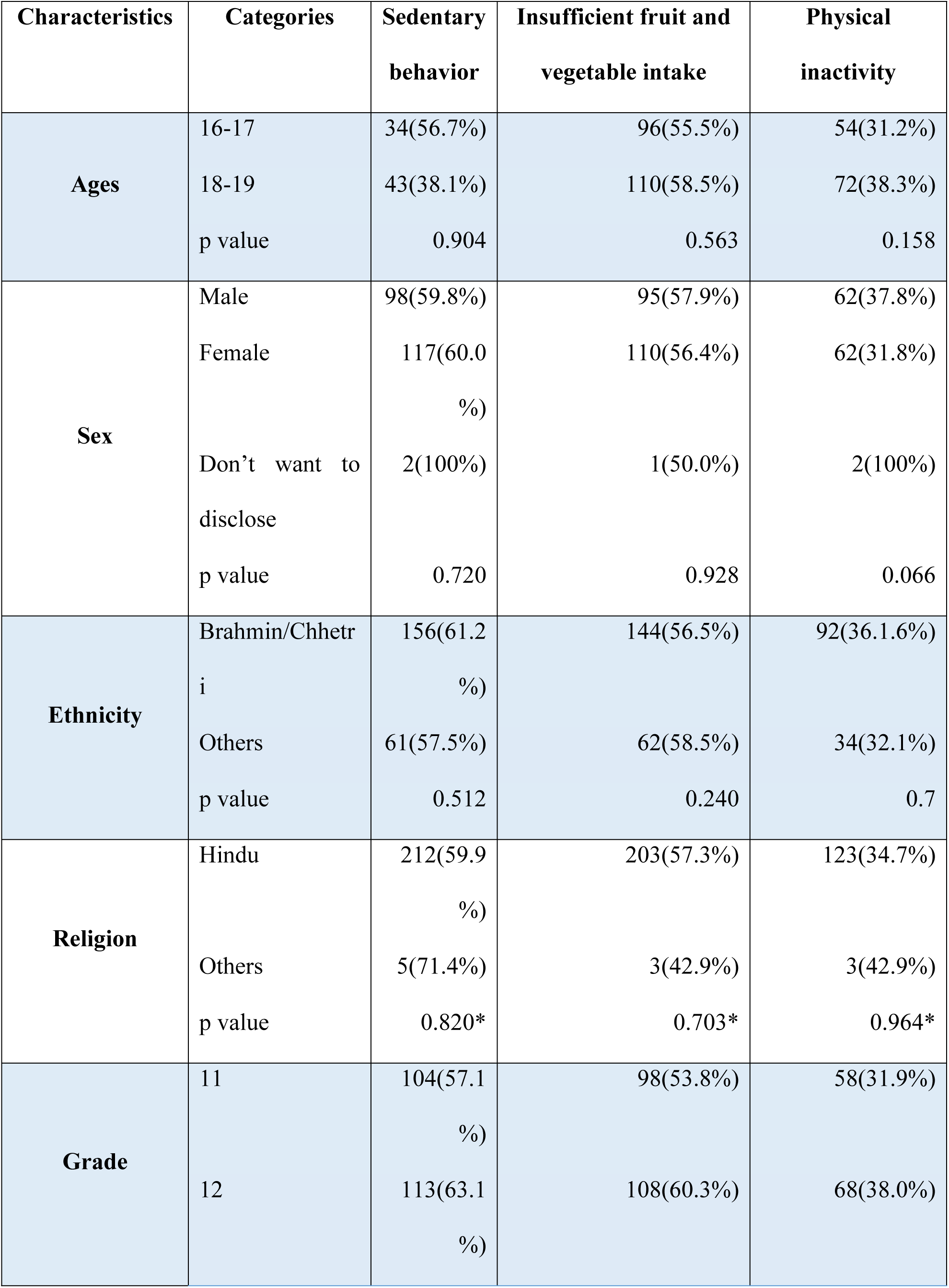
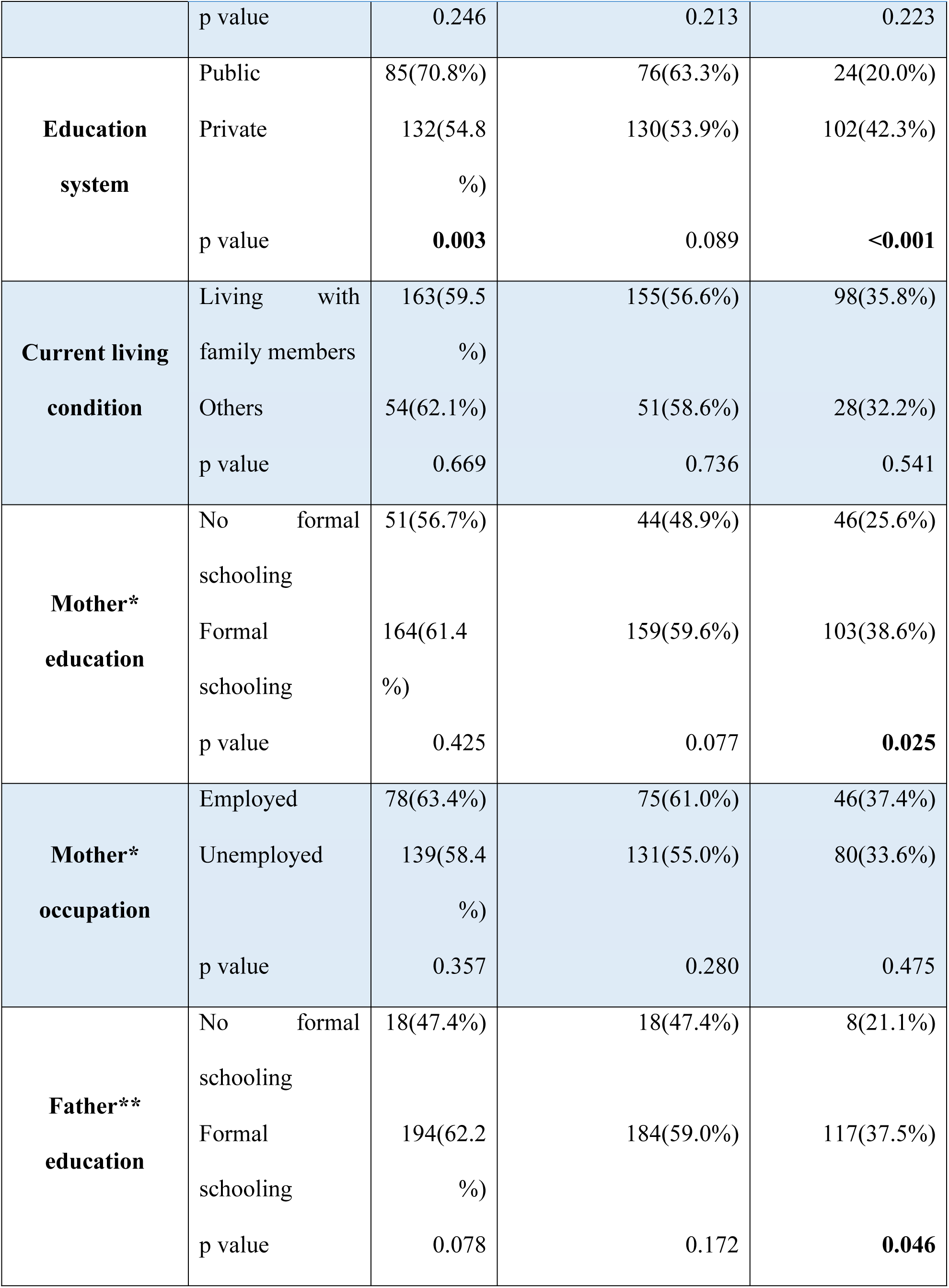
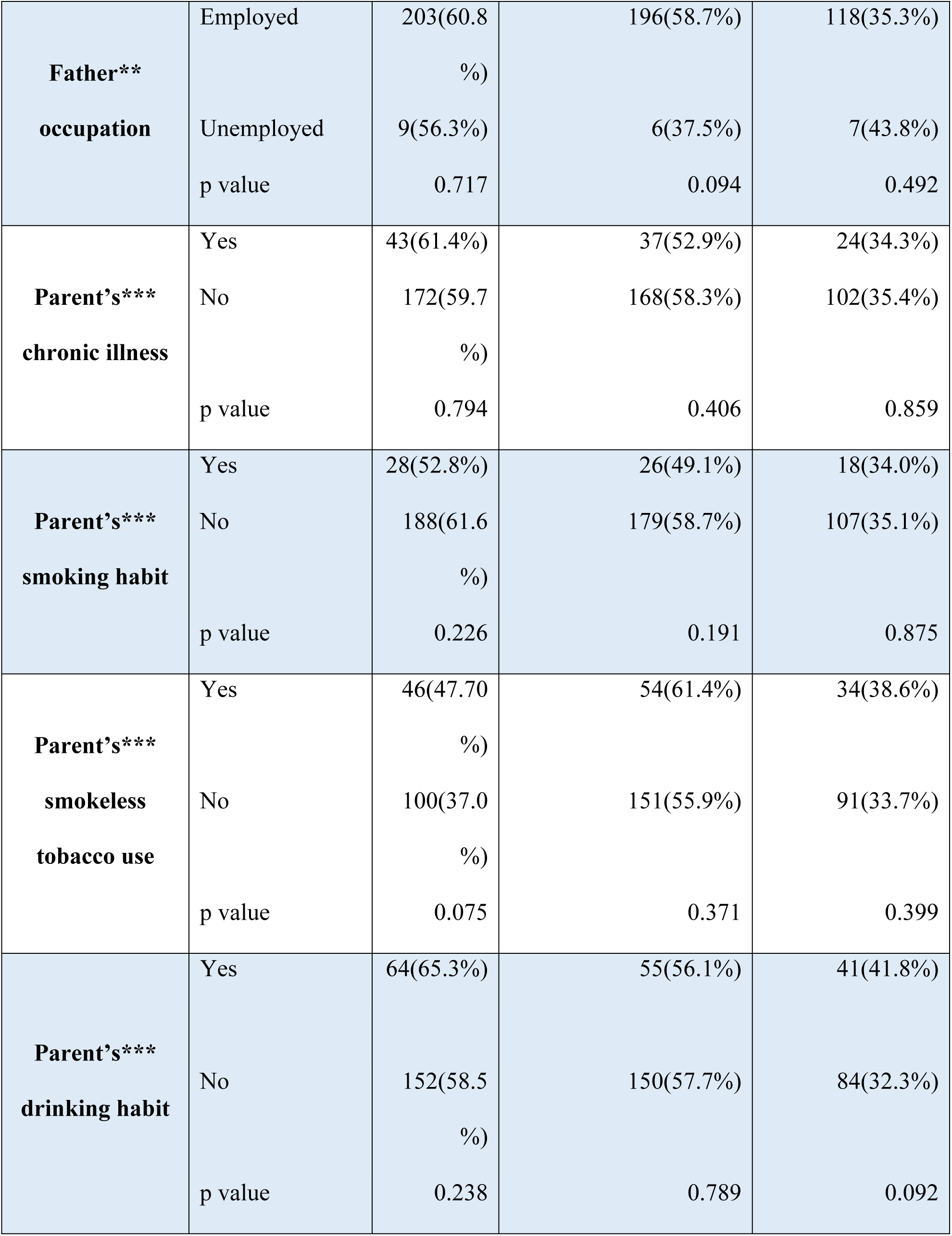
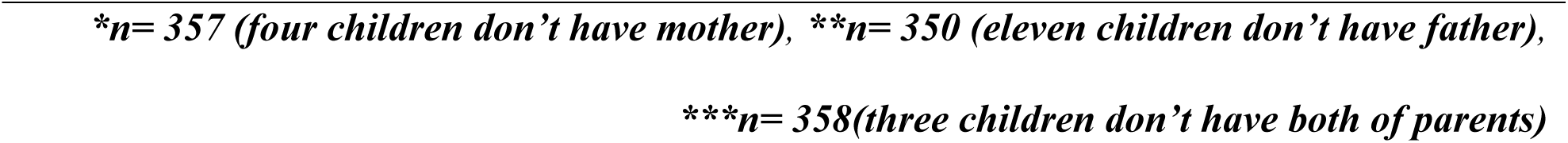
Association of sedentary behaviour, insufficient fruit and vegetable intake, and physical inactivity.

| Characteristics | Categories | Sedentary behavior | Insufficient fruit and vegetable intake | Physical inactivity |
| --- | --- | --- | --- | --- |
| Ages | 16-17 | 34(56.7%) | 96(55.5%) | 54(31.2%) |
|  | 18-19 | 43(38.1%) | 110(58.5%) | 72(38.3%) |
|  | p value | 0.904 | 0.563 | 0.158 |
| Sex | Male | 98(59.8%) | 95(57.9%) | 62(37.8%) |
|  | Female | 117(60.0%) | 110(56.4%) | 62(31.8%) |
|  | Don't want to disclose | 2(100%) | 1(50.0%) | 2(100%) |
|  | p value | 0.720 | 0.928 | 0.066 |
| Ethnicity | Brahmin/Chhetri | 156(61.2%) | 144(56.5%) | 92(36.1.6%) |
|  | Others | 61(57.5%) | 62(58.5%) | 34(32.1%) |
|  | p value | 0.512 | 0.240 | 0.7 |
| Religion | Hindu | 212(59.9%) | 203(57.3%) | 123(34.7%) |
|  | Others | 5(71.4%) | 3(42.9%) | 3(42.9%) |
|  | p value | 0.820* | 0.703* | 0.964* |
| Grade | 11 | 104(57.1%) | 98(53.8%) | 58(31.9%) |
|  | 12 | 113(63.1%) | 108(60.3%) | 68(38.0%) |
|  | p value | 0.246 | 0.213 | 0.223 |
| **Education system** | Public | 85(70.8%) | 76(63.3%) | 24(20.0%) |
|  | Private | 132(54.8%) | 130(53.9%) | 102(42.3%) |
|  | p value | **0.003** | 0.089 | **<0.001** |
| **Current living condition** | Living with family members | 163(59.5%) | 155(56.6%) | 98(35.8%) |
|  | Others | 54(62.1%) | 51(58.6%) | 28(32.2%) |
|  | p value | 0.669 | 0.736 | 0.541 |
| **Mother\* education** | No formal schooling | 51(56.7%) | 44(48.9%) | 46(25.6%) |
|  | Formal schooling | 164(61.4%) | 159(59.6%) | 103(38.6%) |
|  | p value | 0.425 | 0.077 | **0.025** |
| **Mother\* occupation** | Employed | 78(63.4%) | 75(61.0%) | 46(37.4%) |
|  | Unemployed | 139(58.4%) | 131(55.0%) | 80(33.6%) |
|  | p value | 0.357 | 0.280 | 0.475 |
| **Father\*\* education** | No formal schooling | 18(47.4%) | 18(47.4%) | 8(21.1%) |
|  | Formal schooling | 194(62.2%) | 184(59.0%) | 117(37.5%) |
|  | p value | 0.078 | 0.172 | **0.046** |
| **Father\*\***<br>**occupation** | Employed | 203(60.8<br>%) | 196(58.7%) | 118(35.3%) |
|  | Unemployed | 9(56.3%) | 6(37.5%) | 7(43.8%) |
|  | p value | 0.717 | 0.094 | 0.492 |
| **Parent's\*\*\***<br>**chronic illness** | Yes | 43(61.4%) | 37(52.9%) | 24(34.3%) |
|  | No | 172(59.7<br>%) | 168(58.3%) | 102(35.4%) |
|  | p value | 0.794 | 0.406 | 0.859 |
| **Parent's\*\*\***<br>**smoking habit** | Yes | 28(52.8%) | 26(49.1%) | 18(34.0%) |
|  | No | 188(61.6<br>%) | 179(58.7%) | 107(35.1%) |
|  | p value | 0.226 | 0.191 | 0.875 |
| **Parent's\*\*\***<br>**smokeless**<br>**tobacco use** | Yes | 46(47.70<br>%) | 54(61.4%) | 34(38.6%) |
|  | No | 100(37.0<br>%) | 151(55.9%) | 91(33.7%) |
|  | p value | 0.075 | 0.371 | 0.399 |
| **Parent's\*\*\***<br>**drinking habit** | Yes | 64(65.3%) | 55(56.1%) | 41(41.8%) |
|  | No | 152(58.5<br>%) | 150(57.7%) | 84(32.3%) |
|  | p value | 0.238 | 0.789 | 0.092 |

*\*n= 357 (four children don't have mother), \*\*n= 350 (eleven children don't have father),*
*\*\*\*n= 358(three children don't have both of parents)*

Likewise, refined vegetable oil use was associated with ethnicity (p=0.014), education system (p=0.001), current living condition (p=0.026), parents’ chronic illness (0.026), parents’ alcohol use (p= 0.003). Current smokers were associated with gender (p=0.001), ethnicity (p=0.012), mother education (p=0.029), parents smoking habit (p=0.002), parents’ smokeless tobacco habit (p=0.001). Current alcohol use was significantly associated with ethnicity (p = 0.05), parent’s smokeless tobacco use (p = 0.017), and parent’s alcohol use (p = 0.022), and current smokeless tobacco use was significantly associated with gender (p =0.001) as presented in table 6.

**Table 6:**
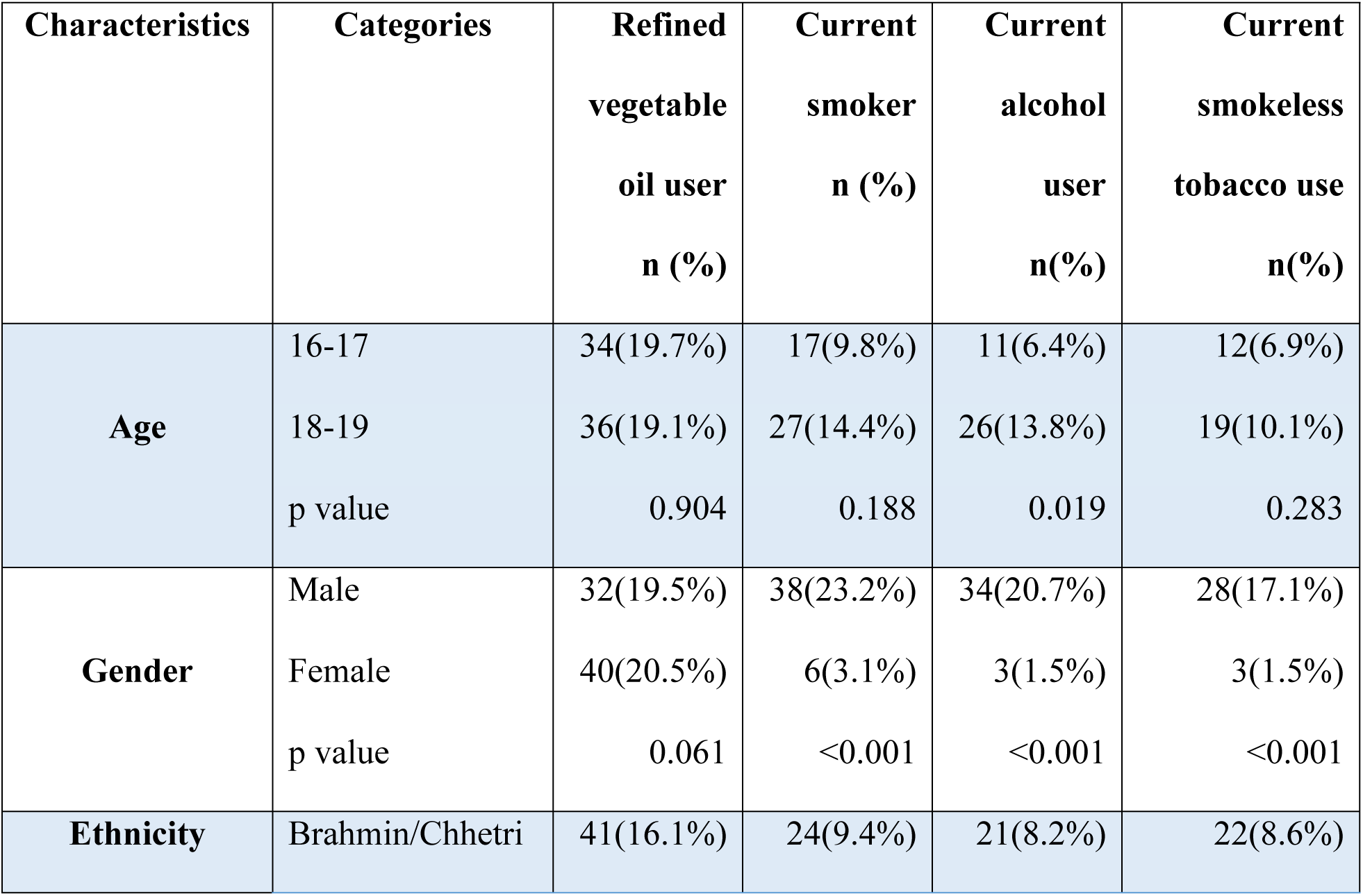
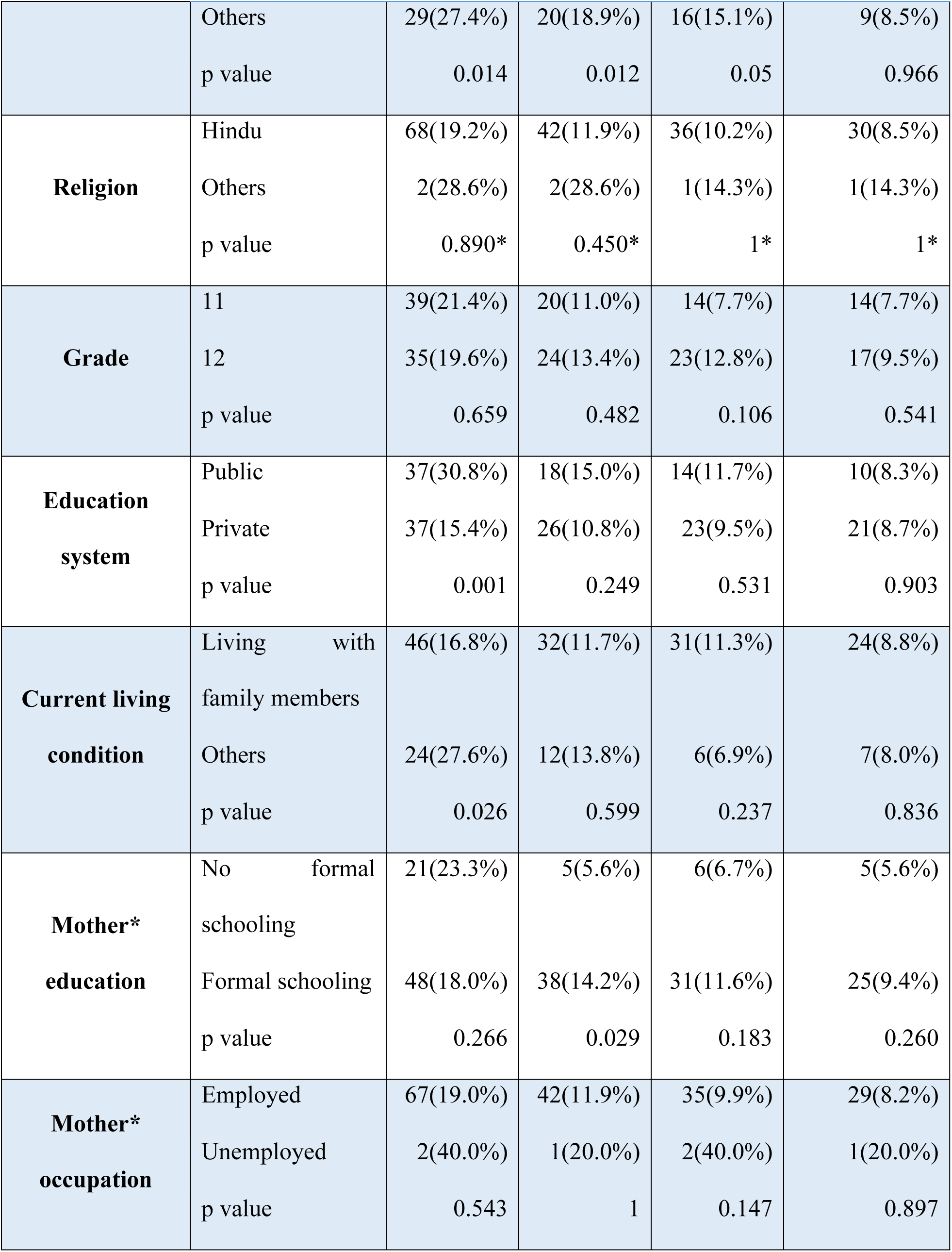
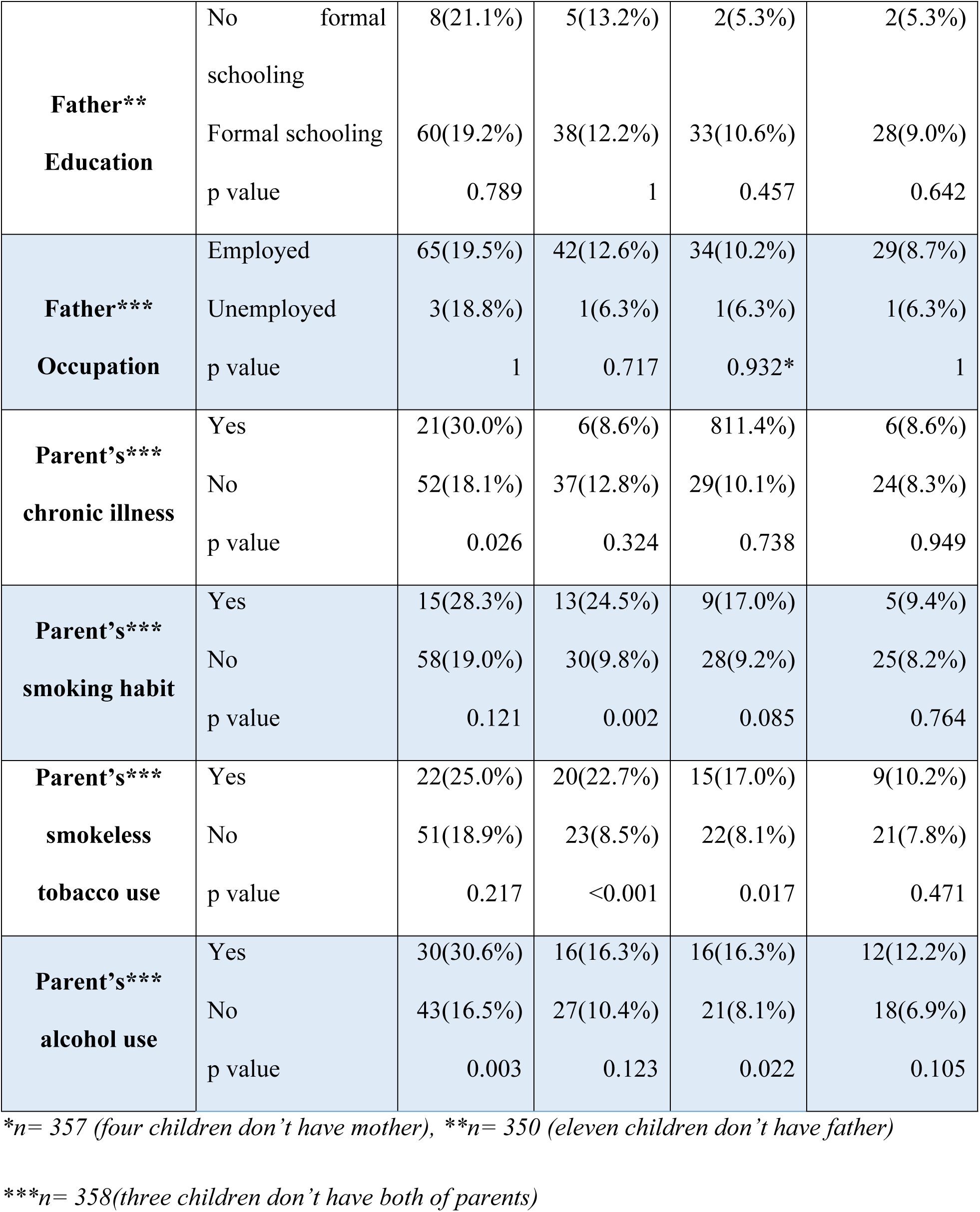
Association of refined vegetable oil use, current tobacco smoking, current alcohol use and current smokeless tobacco use.

| **Characteristics** | **Categories** | **Refined vegetable oil user**<br>**n (%)** | **Current smoker**<br>**n (%)** | **Current alcohol user**<br>**n(%)** | **Current smokeless tobacco use**<br>**n(%)** |
| --- | --- | --- | --- | --- | --- |
| **Age** | 16-17 | 34(19.7%) | 17(9.8%) | 11(6.4%) | 12(6.9%) |
|  | 18-19 | 36(19.1%) | 27(14.4%) | 26(13.8%) | 19(10.1%) |
|  | p value | 0.904 | 0.188 | 0.019 | 0.283 |
| **Gender** | Male | 32(19.5%) | 38(23.2%) | 34(20.7%) | 28(17.1%) |
|  | Female | 40(20.5%) | 6(3.1%) | 3(1.5%) | 3(1.5%) |
|  | p value | 0.061 | <0.001 | <0.001 | <0.001 |
| **Ethnicity** | Brahmin/Chhetri | 41(16.1%) | 24(9.4%) | 21(8.2%) | 22(8.6%) |
|  | Others | 29(27.4%) | 20(18.9%) | 16(15.1%) | 9(8.5%) |
|  | p value | 0.014 | 0.012 | 0.05 | 0.966 |
| **Religion** | Hindu | 68(19.2%) | 42(11.9%) | 36(10.2%) | 30(8.5%) |
|  | Others | 2(28.6%) | 2(28.6%) | 1(14.3%) | 1(14.3%) |
|  | p value | 0.890* | 0.450* | 1* | 1* |
| **Grade** | 11 | 39(21.4%) | 20(11.0%) | 14(7.7%) | 14(7.7%) |
|  | 12 | 35(19.6%) | 24(13.4%) | 23(12.8%) | 17(9.5%) |
|  | p value | 0.659 | 0.482 | 0.106 | 0.541 |
| **Education system** | Public | 37(30.8%) | 18(15.0%) | 14(11.7%) | 10(8.3%) |
|  | Private | 37(15.4%) | 26(10.8%) | 23(9.5%) | 21(8.7%) |
|  | p value | 0.001 | 0.249 | 0.531 | 0.903 |
| **Current living condition** | Living with family members | 46(16.8%) | 32(11.7%) | 31(11.3%) | 24(8.8%) |
|  | Others | 24(27.6%) | 12(13.8%) | 6(6.9%) | 7(8.0%) |
|  | p value | 0.026 | 0.599 | 0.237 | 0.836 |
| **Mother\* education** | No formal schooling | 21(23.3%) | 5(5.6%) | 6(6.7%) | 5(5.6%) |
|  | Formal schooling | 48(18.0%) | 38(14.2%) | 31(11.6%) | 25(9.4%) |
|  | p value | 0.266 | 0.029 | 0.183 | 0.260 |
| **Mother\* occupation** | Employed | 67(19.0%) | 42(11.9%) | 35(9.9%) | 29(8.2%) |
|  | Unemployed | 2(40.0%) | 1(20.0%) | 2(40.0%) | 1(20.0%) |
|  | p value | 0.543 | 1 | 0.147 | 0.897 |
| **Father\*\***<br>**Education** | No formal schooling | 8(21.1%) | 5(13.2%) | 2(5.3%) | 2(5.3%) |
|  | Formal schooling | 60(19.2%) | 38(12.2%) | 33(10.6%) | 28(9.0%) |
|  | p value | 0.789 | 1 | 0.457 | 0.642 |
| **Father\*\*\***<br>**Occupation** | Employed | 65(19.5%) | 42(12.6%) | 34(10.2%) | 29(8.7%) |
|  | Unemployed | 3(18.8%) | 1(6.3%) | 1(6.3%) | 1(6.3%) |
|  | p value | 1 | 0.717 | 0.932* | 1 |
| **Parent's\*\*\***<br>**chronic illness** | Yes | 21(30.0%) | 6(8.6%) | 8(11.4%) | 6(8.6%) |
|  | No | 52(18.1%) | 37(12.8%) | 29(10.1%) | 24(8.3%) |
|  | p value | 0.026 | 0.324 | 0.738 | 0.949 |
| **Parent's\*\*\***<br>**smoking habit** | Yes | 15(28.3%) | 13(24.5%) | 9(17.0%) | 5(9.4%) |
|  | No | 58(19.0%) | 30(9.8%) | 28(9.2%) | 25(8.2%) |
|  | p value | 0.121 | 0.002 | 0.085 | 0.764 |
| **Parent's\*\*\***<br>**smokeless**<br>**tobacco use** | Yes | 22(25.0%) | 20(22.7%) | 15(17.0%) | 9(10.2%) |
|  | No | 51(18.9%) | 23(8.5%) | 22(8.1%) | 21(7.8%) |
|  | p value | 0.217 | <0.001 | 0.017 | 0.471 |
| **Parent's\*\*\***<br>**alcohol use** | Yes | 30(30.6%) | 16(16.3%) | 16(16.3%) | 12(12.2%) |
|  | No | 43(16.5%) | 27(10.4%) | 21(8.1%) | 18(6.9%) |
|  | p value | 0.003 | 0.123 | 0.022 | 0.105 |

\*n= 357 (four children don't have mother), \*\*n= 350 (eleven children don't have father)
\*\*\*n= 358(three children don't have both of parents)

## DISCUSSION

Our study demonstrated the high burden of behavioural risk factors among school going adolescents was disproportionately distributed with male children and students from lower grades, from lower social ethnic groups (considered), private school, staying away from family. In addition, students whose parents were consuming alcohol, tobacco products and history of chronic illness had higher rates of behavioural risk factors.

### Insufficient physical activity and Sedentary behavior

Insufficient physical activity is the 4th leading risk factor for mortality (10). People with insufficient physical activity have a 20% to 30% increased risk mortality compared to those who engage moderate intensity physical activity (10). Our study showed the insufficient physical activity among 35% of adolescents which is higher than insufficient levels of physical activity (10.8%) among adolescents (15–17) of Nepal as reported in STEPS survey 2019 (7), and study conducted in Bangladesh (8%) among adolescent (9). However, our finding is comparable (38%) with the study from Western part of Nepal (11) and very lower (85%) than the Global School Based Student health Survey 2015 findings of Nepal (6) The reason of such discrepancies could be due to recent upgrading of Tulsipur to sub-metropolitan city (urban) from municipality and other socio-cultural differences. In addition, a higher percentage of students were engaged in household work and walking was the most preferred method for going school in this study. The previous study at Lekhnath Municipality showed significant association of physical inactivity with study grades (11). However, our study found a significant association of physical inactivity with education system, mother education and father education. Children of parent with upper secondary education or above have shown significant association with increased physical activity (12). There is evidence that prolonged periods sedentary time was associated with increased risk of CVD mortality, and each additional sedentary time was associated with an increased CVD mortality risk (13). Our study also reported a higher (60%) sedentary behavior among adolescents which is associated with education system. A study from Brazil reported prevalence of sedentary behavior among adolescent was 58.1% (14). The factors such as public schooling, and no formal parental education are indirect indicators of lower socio-economic status and several studies have indicated increased income results in lower physical activities (15). Many other studies have reported that sedentary behavior is more prevalent in children from urban area (16–18). It might be due to less availability of playing ground and space for physical activities and high opportunities of sedentary life style such as access to transportation and other high scree time (video games… etc.).

### Low fruit and vegetable intake and Calorie drink consumption

Low fruit and vegetable intake is associated with an increased risk of stroke by 11% and ischemic heart disease by 31% (19). WHO estimates that 2.7 million lives could be saved each year if fruits and vegetables are consumed adequately (19). More than half (57.1 %) of adolescents were taking insufficient fruit and vegetable in our study which is similar (58 %) to the findings of the study conducted in Western part of Nepal (11). However, our findings is lower than findings of Bangladesh (98%) among adolescents aged 12-18 (9). The difference in Bangladesh could be due to differential inclusion criteria of both rural and urban adolescents of age group 12-18 in their study (9). There were not any associated socio-demographic or parental factors identified in our study.

Our study reported the high consumption (84%) of the calorie drink among adolescents which is corroborates (83.1%) to study from the Western part of Nepal (11). In contrast, our findings is higher than that of nation-wide survey Global School Based Student Health Survey Nepal 2015 (33.3%) and the study conducted in India (44.8%) (6, 20). These differences could be due to focus of our study to an age group from urban area. In addition, it could be due to rapid unplanned urbanization of city replacing access to fruit and vegetable intake by other easily available fast and junk foods and calorie drinks among adolescents. However, we did not find any association with calorie drink consumption.

### Added salt intake and consumption of processed food high in salt

High salt intake is one of the most important determinants of high blood pressure and increased risk of cardiovascular disease. The prevalence of added salt intake (always or often) was found 15.5% in our study which is lower (66.9 %) than that of study conducted in Bangladesh (9). These discrepancies could be due to differential study population and food preference practices among two countries. In addition, mother education was associated (OR 1.8) with added salt intake in our study. Like most of the South Asia countries, mothers are the one who mostly prepared food for their kids in Nepal. And due to illiteracy of mothers on harmful health impacts of added extra table salt, adolescents might be exposed to such risk factors. It demands education to the kids including their parents is very important to tackle such escalating CVD risk factors among adolescents.

In addition, our study reported 33.0% consumption of processed food high in salt which is lower (75%) the findings of Global School Health Based Health Survey and then a study conducted in India (49.2%) among school children (20,21). The differences of prevalence could be due to differential presentation of finding as reported in those study as consumption for at least once a day per week and our study reported basis of always/daily or often/most days a week consumption of processed food high in salt. In addition, this could be due to socio-economic background associated with consumption of processed food high in salt due to available of more pocket money among school going adolescents and broadening of health knowledge according to grades.

### Smoking Tobacco use and Smokeless tobacco use

Acute cardiovascular effects, like those caused by cigarette smoking, are seen with the use of smokeless tobacco. Each year, about 5 million people die as a result of direct tobacco smoking around the world, with many of these deaths occurring prematurely where tobacco use is responsible for 10% of all deaths from cardiovascular diseases (22). The prevalence of current smoking tobacco use found in this study is higher than the prevalence found in Global School Health Survey and a study conducted in Bangladesh (9, 21). Current smoking tobacco use was found significantly associated with gender (OR=0.1015), ethnicity (OR=2.238), mother education (OR=2.821), parent’s smoking habit (0.336) and parent’s smokeless tobacco use(OR=0.317). The reasons behind association could be due imitating of parental behavior by adolescent as our study found the association with parents smoking habits. It is found that children from highest wealth quintile are likely to be smokers than from lower health quintile, association with no formal mothers education in our study can be linked with lower socio-economic status (23).

In addition, current smokeless tobacco user in our study were similar to the findings of Global School Health Based Survey and higher than study of Bangladesh (9, 21). The relative risk of being current user of smokeless tobacco among males was 11 times higher than female in our study which is comparable to previous studies (23). The reason could be stereotypical society in Nepal accepting and promoting the tobacco consumption among males. In addition, it could be due to lack of demand reduction provisions of tobacco adopted by study area and easily accessibility of cigarette to adolescents in market. Importantly, it could be due to poor implication of tobacco demand reduction policy in Nepal (24) leading to easily accessibility of tobacco products to minors in the market. Our study had included some independent variables such as education system, current living condition and socio-demographic, medical and behavioral characteristics of parents which had not been included in most of previous literature.

### Harmful use of alcohol

Worldwide, 3 million deaths every year result from harmful use of alcohol which is attributed to 5.3% of all deaths (25). There is evidence that alcohol consumption is positively associated with risk of atrial fibrillation, heart failure, and hemorrhagic stroke (26–28). Overall 10.2% of adolescents were found to have drinking alcohol currently which is higher than the findings of Global School Health Based Survey (6). However, it is lower than the study findings from Western Nepal (11). The difference could be due to easily accessibility of various alcoholic drinks in the market (6). This study showed significant association with age (OR=0.401) gender (OR=16.738), ethnicity (OR=0.505), parent’s smokeless tobacco use (OR=2.314) and parent’s alcohol use (2.221). The reason for association with age, gender and parental habits could be due to cultural background, gender stereotypes and influence of parent’s behavior on children. This is an area of future research and intervention planning focusing on both parents and children.

## STRENGTHS AND LIMITATIONS

Our study studied the relation of some crucial factors such as the education system, current living conditions and socio-demographic, medical and behavioral characteristics of parents with CVD risk, which was focused by limited previous studies to our knowledge.

Several factors might affect the findings of this study. First, the results represent the prevalence of adolescents aged 16-19 years in Tulsipur Sub-Metropolitan City which cannot be generalized in a general adolescent population of the different settings of Nepal, such as rural areas. A self-administered questionnaire was used to assess the risk factors that might lead to self-reported and recall bias. Collecting data in a group setting might have been subjected to information bias which was addressed by not allowing more than 15 participants with adequate spacing. However, we have used standardized questions with appropriate probing. In addition, enough clarification of questions was provided before data collection to overcome those biases.

## CONCLUSION

This study provided evidence that most behavioral cardiovascular disease risk factors were disproportionately distributed among different subgroups of adolescents, such as males, underprivileged minority ethnic groups, parental history of CVDs and parental education. A high prevalence of behavioral cardiovascular disease risk factors among adolescents may result in the emergence of CVD in their adult age or later stage. This would be alarming in future which needs high-cost interventions. Thus, there is an urgent need for high-risk group intervention, including both parents and children. In addition, adequate monitoring of the implementation of already developed policies, such as banning smoking and alcoholic products for adolescents and discouraging the marketing of fast/processed foods with salt and high-caloric drink at the municipal level (local government) is urgently needed.

## Data Availability

The authors can share datas without reveling the information regarding research participants when requested.

## GENERALIBILITY

Tulsipur sub-metropolitan city being situated in inner terai region having characteristics of both terai and hilly region and with unplanned rapid urbanization it has a mixed feature of urban and rural area. This study has adequate sample size can be generalized for urban, rural, hilly and plain settings of LMICs.

